# Choroid Plexus calcification correlates with cortical microglial activation in humans: a multimodal PET, CT, MRI study

**DOI:** 10.1101/2022.11.14.22282263

**Authors:** Tracy Butler, X. Hugh Wang, Gloria C. Chiang, Yi Li, Liangdong Zhou, Ke Xi, Nimmi Wickramasuriya, Emily Tanzi, Edward Spector, Ilker Ozsahin, Xiangling Mao, Q. Ray Razlighi, Edward K. Fung, Jonathan P. Dyke, Thomas R. Maloney, Ajay Gupta, Ashish Raj, Dikoma C. Shungu, P. David Mozley, Henry Rusinek, Lidia Glodzik

## Abstract

**Background:** Choroid plexus (CP) within brain ventricles is well-known to produce CSF. Additional important CP functions are now recognized including critical modulation of inflammation. Recent MRI studies have demonstrated CP enlargement in human diseases including Multiple Sclerosis and Alzheimer’s Disease, and in association with neuroinflammation measured using translocator protein (TSPO) PET. The basis of MRI-visible CP enlargement is unknown.

**Purpose:** Based on tissue studies demonstrating CP calcification as a common pathology associated with aging and disease, we hypothesized that previously-unmeasured calcium within CP contributes to MRI-measured CP volume, and may be more specifically associated with neuroinflammation.

**Materials and Methods:** We performed a retrospective analysis of PET-CT studies performed between 2013-2019 on a single scanner using the TSPO radiotracer 11C-PK11195. Subjects included controls (n=43) and patients diagnosed with several non-inflammatory neuropsychiatric conditions (n=46.) Cortical inflammation / microglial activation was quantified as non-displaceable Binding Potential (BPnd.) CP and ventricle volume were measured using Freesurfer. CP calcium was measured semi-manually via tracing of low-dose CT acquired with PET and automatically using a new CT/MRI method. The contribution of CP calcium, CP overall volume, ventricle volume, subject age, sex and diagnosis to BPnd was assessed using linear regression.

**Results:** 89 subjects (mean age 54+/-7 years; 52 men) were included. Fully-automated CP calcium quantification was accurate (ICC with semi-manual tracing = .98.) The significant predictors of cortical neuroinflammation were subject age (p=.002) and CP calcium volume (p=.041), but not ventricle or CP volume.

**Conclusion:** CP calcium volume can be accurately measured using low-dose CT acquired routinely with PET-CT. CP calcification – but not CP overall volume – was associated with cortical inflammation. Unmeasured CP calcification may be relevant to recent reports of CP enlargement in human inflammatory and other diseases. CP calcification may be a specific and relatively easily-acquired biomarker for neuroinflammation and CP pathology.

**Key Results:**

- Choroid plexus (CP) calcification volume can be reliably quantified using semi-manual tracing on low-dose CT acquired with PET-CT, and fully automatically using our new, accurate (ICC with semi-manual tracing = .98) CT/MRI method.
- CP calcification and age –but not overall CP volume– significantly predicted 11C-PK11195 PET-measured cortical neuroinflammation in 89 subjects.
- CP calcification is a relatively easily-assessed, previously-overlooked potential biomarker for neuroinflammation and CP pathology.

## INTRODUCTION

The choroid plexus (CP), a highly vascularized filamentous structure within the brain ventricular system, constitutes the blood-CSF barrier and is well known to produce CSF. Recently, additional, important CP functions have been recognized including clearance of toxins ^1, 2^, sleep-wake regulation ^3^, secretion of hormones and growth factors ^4^, support of neurogenesis ^5, 6^, and regulation of inflammation. The role of CP in inflammation is especially critical: CP both initiates and modulates neuroinflammation, and CP dysfunction is considered highly relevant to the pathophysiology of multiple sclerosis (MS) ^7-12^.

Unlike virtually all brain parenchymal structures, which atrophy in association with aging and dysfunction, CP *enlargement* appears to be pathological. Recent MRI studies have demonstrated larger CP volume in association with normal aging ^13^ and in patients with neuropsychiatric disorders including Alzheimer’s Disease ^14, 15^, depression ^16^, psychosis ^17^, stroke^18^, and multiple sclerosis ^11, 12^. The basis of CP enlargement remains uncertain, but based on autopsy and animal studies, has been posited to relate to CP basement membrane thickening and fibrous stroma expansion with protein, inflammatory cells and deposition of metal, in particular calcium ^19-23^. While most of these CP components are detectable and quantifiable only through direct examination of tissue, assessment of calcium using *in vivo* neuroimaging is relatively simple because of its vastly higher photon attenuation compared to water and brain, making it easily visible on Computed Tomography (CT) ^24^. For decades, CP calcification has been recognized as a normal phenomenon, with gradually increasing calcification noted with increasing age ^25^. Markedly increased calcification of CP, basal ganglia, and other brain structures in association with neuropsychiatric decline constitutes the still poorly understood and underdiagnosed syndrome of Fahr disease ^26^. CP calcification has received less research attention in recent years in large part because MRI, with its submillimeter spatial resolution and lack of radiation, has supplanted CT as the modality of choice for human neuroimaging research studies, and the variable signal of calcium on typical MR sequences makes calcium quantification on MR challenging ^27^. To our knowledge, decades-old recognition of increasing CP calcification with aging and certain diseases has not been linked to more recent understanding that CP has important roles beyond CSF production, in particular, critical regulation of neuroinflammation ^7-12^. We therefore took a multimodal imaging approach, combining CT, MRI and Positron Emission Tomography (PET) with 11C-PK11195, a radiotracer sensitive to the translocator protein (TSPO) expressed by activated microglia ^28^ to measure the relationship between CP calcium and cortical neuroinflammation in a large group of human subjects. We developed an automated method for measuring CP calcium volume using low-dose CT (routinely acquired during PET-CT studies) and MRI, which we validated against semi-manual tracing. We hypothesized that the volume of calcium within CP would be an independent predictor of PET-measured neuroinflammation.

## METHODS

### Subjects

Subjects (n=89) who underwent 11C-PK11195 (PK) PET for several different studies conducted on a single Siemens Biograph mCT PET-CT scanner between 2013 and 2019 were included in this analysis. Subjects were either healthy controls (n=43) or had been diagnosed with Chronic Fatigue Syndrome (CFS; n=17), Gulf War Illness (GWI; n=12) or Parkinson’s Disease without dementia (PD; n=17.) Control subjects were free of significant medical, neurologic and psychiatric disease. All subjects provided informed consent to participate in a disease-focused research study and for their data to be included in a registry for future analyses such as this one. Sex differences in hypothalamic inflammation in healthy controls from this dataset have been previously reported ^29^.

### Image Acquisition

PET images were acquired over one hour starting at the time of injection of ∼550 MBq of 11C-PK11195. List-mode emission data were reconstructed using OSEM method with four iterations and 21 ordered subsets, attenuation and scatter corrected using attenuation map derived from CT, and binned into 22 frames.

A low-dose CT image was acquired immediately prior to PET acquisition. CT parameters included tube voltage: 120 kVp, tube current: 35 mA, pitch factor: 1.5 and voxel size: 0.5×0.5×1.5mm^3^.

Three-dimensional volumetric T1 weighted BRAVO/MPRAGE MR images (1.2 × 1.2 × 1.2 mm^3^ isotropic voxels) were acquired using a GE Signa or Siemens Skyra 3T scanner.

### PET Image Processing

PET frames were motion corrected using MCFLIRT^30^ within FSL ^31^. Nondisplaceable Binding Potential (BPnd) images reflecting the concentration of translocator protein (TSPO) expressed by activated microglia, irrespective of tracer delivery/blood flow, were generated from dynamic PET using a multilinear reference tissue model ^32^ implemented in the freely available software package FireVoxel (https://firevoxel.org.) The reference tissue time-activity curve was identified via optimized supervised cluster analysis ^33^ which is considered the method of choice for analyzing PK PET scans ^34^.

### MR Image Processing

Freesurfer v7^35^ was used to segment MR images for three purposes: to define the bilateral cortical region of interest (ROI) for quantifying PET signal; to segment bilateral lateral ventricles for the automated calcium quantification method described below; and to quantify CP and lateral ventricle volume to include as predictors of cortical inflammation. For this latter purpose, CP and ventricular volumes were divided by each subject’s whole brain volume, calculated using SPM 12, to account for head size. To quantify inflammation (average BPnd) over the cortical ROI, each subject’s T1W MRI was linearly co-registered to his/her summed PET image with rigid body transformation in FSL ^31^. ROIs were transformed to PET space with the inverse transformation matrix from the co-registration step and eroded 2mm in plane to minimize partial volume effects.

### Semi-manual quantification of CP calcium on CT

Using FireVoxel, CT images were windowed at width=100 Hounsfield Units (HU) and level=50 HU to clearly visualize CP calcifications, which typically have signal greater than 100 HU.^25^ Three dimensional ROIs of CP calcification within all horns of bilateral lateral ventricles were drawn using an electronic paintbrush on contiguous axial images. ROIs were then thresholded at 60 HU to exclude any non-calcified voxels. Because neither brain parenchyma nor CSF adjacent to CP approximates the high signal of calcium, manual tracing could be done quickly, and any mistakenly-included non-calcified voxels removed automatically by thresholding. Semi-manual tracing was performed by one rater on a random subset of scans (n=44.) To assess inter-rater reliability, a second rater traced 10 of these scans.

### Automated quantification of CP calcium using MRI and CT

T1 MR and Freesurfer segmentations were co-registered to CT using Advanced Normalization Tools (ANT’s)^36^ rigid transformation. An ROI mask consisting of bilateral CP and the posterior portion of bilateral lateral ventricles (posterior to the most anterior extent of CP) was generated. Excluding the most anterior portion of this mask prevented erroneous misidentification of parenchymal voxels as calcified, and was found to improve correlation with manual tracing.

Voxels with HU>100 within this combined CP/posterior LV mask were used as seeds to build a weighted image based on the intensity difference. This weighted image was segmented to background and foreground using fast marching method.^37^ Clusters from the segmentation were then divided into two groups with kmeans methods^38^ and the one with higher HU value selected as the calcified CP voxels. **Figure 1** shows an example of CP semi-manual and automated segmentation.

**Figure 1.**
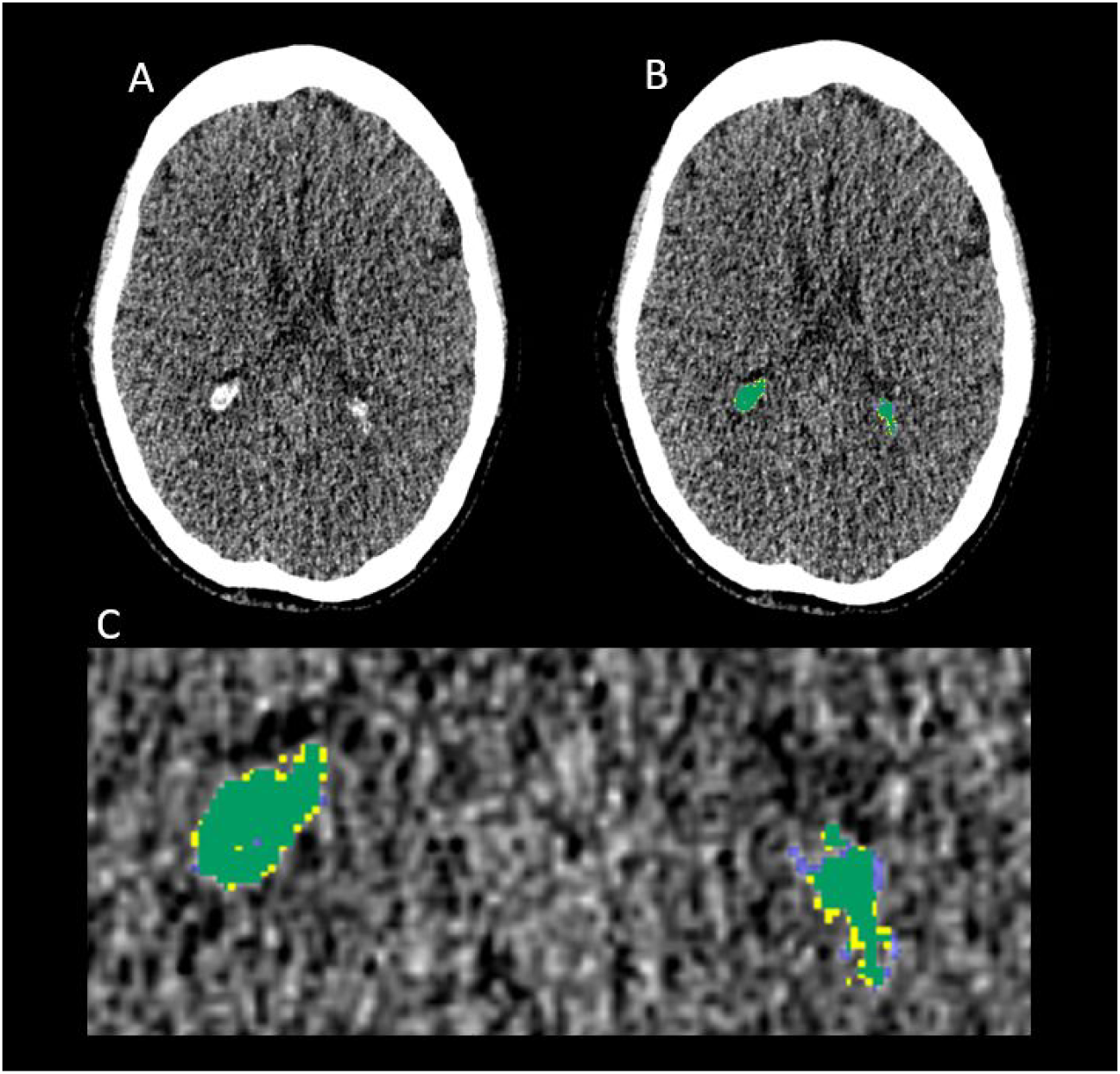
Example of CP calcification segmentation. **A.** axial slice from low-dose CT acquired with PET-CT shows CP calcification in the glomus of the lateral ventricles bilaterally. **B.** Semi-manually traced CP calcification is shown in yellow, fully-automated segmentation in blue and the overlap in green. **C.** Magnified view of B showing close correspondence between semi-manual and automated CP calcification segmentation.

### Statistical Analyses

Analyses were performed in SPSS version 26. All results were considered significant at p<.05. Intraclass correlation coefficient (ICC) was used to assess inter-rater reliability between the two semi-manual CP calcium tracers and agreement between semi-manual and automated CP calcium volumes.

Differences in CP calcium volume, CP overall volume and cortical BPnd between the diagnostic groups and between women and men were assessed using ANCOVA, controlling for age. Multiple linear regression was used to assess the contribution of CP calcium, CP volume, ventricle volume, subject age, diagnosis and sex to the dependent variable of cortical BPnd.

## RESULTS

<u>Subject demographics</u> are presented in **Table 1**.

**Table 1.**
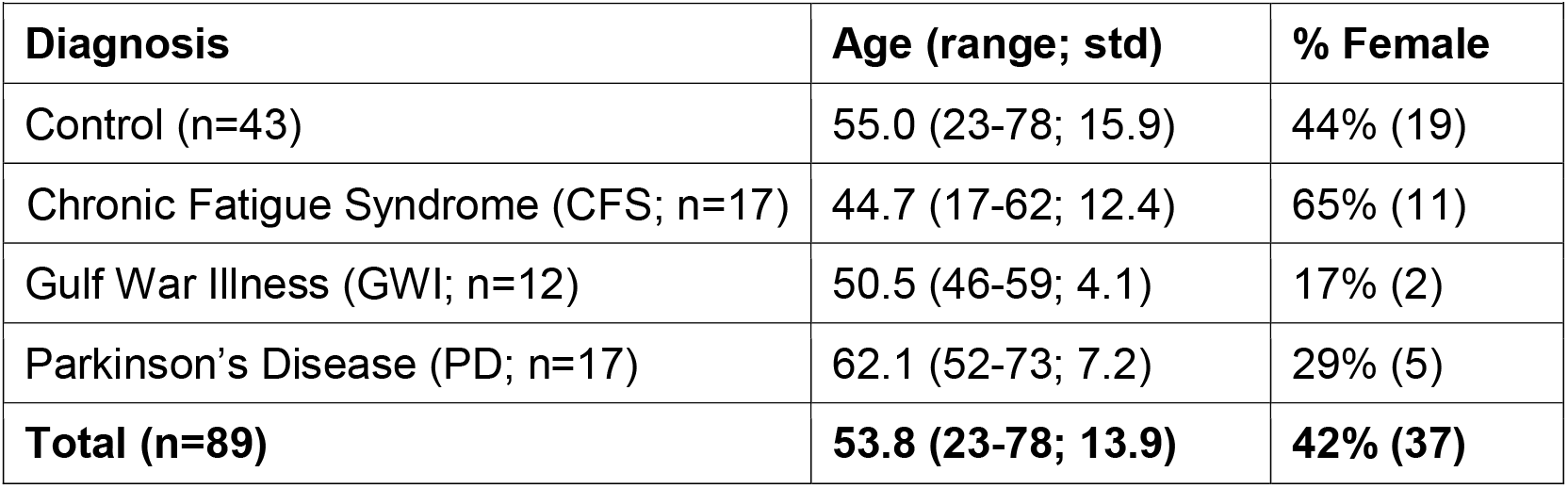
Subject demographics.

| **Table 1.** Subject demographics. |  |  |
| --- | --- | --- |
| **Diagnosis** | **Age (range; std)** | **% Female** |
| Control (n=43) | 55.0 (23-78; 15.9) | 44% (19) |
| Chronic Fatigue Syndrome (CFS; n=17) | 44.7 (17-62; 12.4) | 65% (11) |
| Gulf War Illness (GWI; n=12) | 50.5 (46-59; 4.1) | 17% (2) |
| Parkinson's Disease (PD; n=17) | 62.1 (52-73; 7.2) | 29% (5) |
| **Total (n=89)** | **53.8 (23-78; 13.9)** | **42% (37)** |

### Comparison between semi-manual and automated CP calcium measurement

There was excellent absolute agreement between the two semi-manual tracers (ICC=.990, F(9,9)= 181.58, p<.001) and between semi-manual and automated calcium measurements (ICC= .982, F(43,43)= 71.42, p<.001.) Based on this, for all subsequent analyses, CP calcium refers to the automated value.

### Group and sex Differences

When controlling for age, which differed significantly across diagnostic groups (GWI and CFS subjects were younger than controls and PD subjects) there were no group or sex differences in CP calcium volume, CP overall volume or cortical BPnd (p>.1 for all analyses.)

### Association of CP calcium, CP overall volume, ventricle volume, age, sex and diagnosis with cortical BPnd

As shown in **Table 2**, a multiple regression model (R^2^ = .257, F(6, 82) = 4.72, p < .001) showed that only age and CP calcium volume were significant predictors of cortical BPnd. Results were similar when one subject with high CP calcium volume (1407.6 mm; 5.2 std > mean; shown in **Figure 2**) was excluded from analysis: (R^2^ = .243, F(6, 81) = 4.326, p = .001; CP Calcium Standardized β =.204; p=.05.)

**Table 2.** Linear regression model predicting cortical BPnd.

| **Table 2. Linear regression model predicting cortical BPnd.** |  |  |  |
| --- | --- | --- | --- |
| **Variable** | **Unstandardized $\beta$** | **Standardized $\beta$** | **p-value** |
| CP calcium volume | 2.036E<sup>-5</sup> | .220 | **.041** |
| age | .001 | .364 | **.002** |
| CP volume | -1.075 E<sup>-5</sup> | -.143 | .353 |
| Ventricle volume | 3.351 E<sup>-7</sup> | .137 | .340 |
| sex | .005 | .107 | .284 |
| diagnosis | .001 | .078 | .446 |

**Figure 2.**
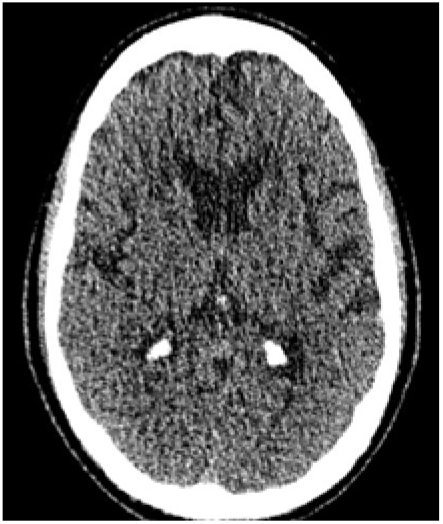
CT showing subject with largest CP calcium volume (1407.6 mm^3^), considered clinically normal.

## DISCUSSION

We show that the volume of calcium within CP – but not CP overall volume – is associated with TSPO PET-measured cortical neuroinflammation, and provide a method for accurately and automatically measuring CP calcium volume using MRI and low-dose CT. These results shed light on the basis of pathological CP enlargement increasingly recognized in association with human inflammatory and neurodegenerative diseases, and suggest the potential for CP calcification to serve as a sensitive, specific and relatively easily-acquired biomarker for both neuroinflammation and CP dysfunction.

### Quantifying CP calcification

We provide a method for fully automated CP calcium volume quantification based on low dose CT routinely acquired during PET-CT and T1 MRI. This method was highly congruent (ICC=.98) with the gold standard of manual tracing.

### Specific association between calcification and activated microglia

The novel result of this study is that CP calcification is independently associated with cortical inflammation when accounting for age, ventricle volume and CP volume, suggesting a possible unique contribution of CP calcification to inflammation. This possibility is supported by recent findings in an animal model of familial brain calcinosis (commonly referred to as Fahr’s disease in humans) directly linking brain calcification to activated microglia.^39^ In this study, activated microglia were *beneficial* in regulating calcification and preventing neurodegeneration. Current findings showing an association between activated microglia and calcification in subjects free from inflammatory disease are broadly consistent with such a homeostatic interplay between microglia and calcification. Calcium in CP has recently been shown to be uniquely dynamic,^40^ and chelation therapy to remove metals from the brain and body is feasible and safe^41^ and in human trials for neurodegenerative disease,^42^ suggesting possible direct therapeutic relevance of CP calcium quantification.

### CP calcium as a sensitive and specific biomarker for neuroinflammation and CP pathology

More broadly, and independent of any direct, mechanistic links between CP calcification and neuroinflammation,^39^ current results provide strong support for use of CP calcification as a biomarker of CP pathology and neuroinflammation, and shed light on the basis of its pathologic enlargement in human diseases. Calcium in CP is found mainly in structures called psammoma bodies, consisting also of collagen, phosphorus and iron.^21^ Psammoma bodies are present only in CP and in certain tumors, and are considered a hallmark of CP aging and dysfunction.^20, 21^ Calcium quantification using *in vivo* neuroimaging would therefore be expected to reflect CP dysfunction and structural abnormalities with greater sensitivity and specificity than CP overall volume measurement, and current results support this: we found that CP calcification, but not CP volume, was a significant predictor of cortical inflammation. It is possible that unmeasured CP calcium can explain prior findings of CP enlargement in human aging and disease^12-18^ including highly relevant recent findings of CP enlargement in association with TSPO PET-measured cortical inflammation.^12, 16^ Additional work is needed to address this issue, including studies linking *in vivo* or *ex vivo* neuroimaging of human CP to direct tissue assessment.

Future studies can benefit from imaging modalities focused specifically on CP assessment. Our retrospective study capitalized on the use of CT acquired during PET to assess a feature of CP (calcification) that has not received recent research attention. In future studies, novel MR sequences can quantify and differentiate calcium from other metals especially iron^41^ with the benefit of greater spatial resolution and no radiation risk to human subjects. CP function and blood flow can be assessed using MRI with ^43^ and without ^44, 45^ gadolinium-based contrast, and modeling of dynamic PET.^46, 47^

### Limitations

This study has several significant limitations. It uses TSPO PET to quantify cortical neuroinflammation indexed by activated microglia. However, TSPO PET is expressed not just by microglia but by several cell types in the brain ^48^ and microglial “activation” is a complex process which cannot be assessed using a probe for a single molecule ^49^. Furthermore, this study used a first generation TSPO radiotracer with inferior signal properties to newer tracers ^28^. However, TSPO PET is the only available method to assess microglial activation *in vivo* in humans, and we apply optimal image processing and analysis methods ^34^.

It can be considered a limitation that our subject group was heterogeneous and included subjects with a variety of disorders, including patients with PD in whom inflammation of substantia nigra in midbrain is pathophysiologically important ^50^. Results were not significant when the subject group was halved to include only normal control subjects. Addressing this limitation, there were no significant differences in CP calcification, CP volume or cortical BPnd between subject groups, we included diagnosis as a covariate in all analyses, and assessment of neuroinflammation was limited to the cortex, which is not considered to be involved in PD without dementia ^50^ or the other included disorders.

When the subject with the highest level of CP calcification, shown in Figure 2, was excluded from analysis, results were weakened (p=.05.) While identification and possible exclusion of outliers is important in any statistical analysis, exclusion of this subject would not reflect the biological reality of wide variability in CP calcification, considered normal in clinical neuroradiology practice.

This correlative cross-sectional imaging study does not allow confirmation of a causal relationship among CP calcification, enlargement and cortical inflammation. Longitudinal imaging studies will be needed to define causality.

## CONCLUSION

Results highlight measurement of CP calcium as a potential biomarker for neuroinflammation and abnormal CP structure and function, and provide an automated method for doing so using CT and MRI. Results contribute to a better understanding of the role of CP in inflammation, aging and human disease, which may have therapeutic implications. CP transplant is technically feasible (much more so than other brain structures) and has been shown to prevent neurodegeneration in animal models.^23^ Because the vast majority of human PET studies are currently performed using PET-CT machines, assessment of the contribution of CP calcification to any type of PET-measured molecular information can be performed with no additional cost or radiation burden to the subject. For example, CP dysfunction has been posited to play an early etiologic role in Alzheimer’s Disease (AD)^1, 2, 14, 15, 19^. Whether CP calcification predicts amyloid beta deposition in AD could be assessed using existing longitudinal neuroimage repositories if the CT portion of the PET-CT were recognized as valuable and made available. We believe current results demonstrate the value of low-dose CT in evaluating CP structural features not easily assessable with MRI and provide critically needed information about the role of CP in human disease.

## Data Availability

All data produced in the present study are available upon reasonable request to the authors

## Funding Acknowledgement

This work was funded by NIH grants R01AG057681, K23NS057579, R01NS105541, R01NS092802, RF1AG062196, R01AG072753 and DOD grant: W81XWH-15-1-0437.

